# Impact of the 2013 WHO guidelines for screening and treatment of cervical precancerous lesions on women’s screening rates, by HIV-status, in East and Southern Africa: A regression discontinuity design analysis

**DOI:** 10.1101/2024.04.05.24305378

**Authors:** David Chipanta, Connie Osborne, Julia Bohlius, Ani Shakarishvilia, Silas Amo-Agyei, Alinane Linda Nyondo-Mipando, Janne Anton Markus Estill, Olivia Keiser

## Abstract

**Introduction:** In 2013, the World Health Organization (WHO) issued guidelines for cervical pre-cancer screening. It recommended screening women aged 30–49, and younger women once they tested HIV-positive. Subsequent WHO guidelines recommended screening women living with HIV (WLHIV) starting at age 25. However, the impact of 2013 guidelines and age to start screening on screening has not been studied.

**Methods:** We used a regression discontinuity design (RDD) analysis of population-based data to assess the impact of the 2013 WHO guidelines on the screening rates for women according to HIV status and age group in Ethiopia, Malawi, Rwanda, Tanzania, Zambia, and Zimbabwe. The outcome was self-reported ever having been screened for cervical pre-cancer between 2008 and 2018. We compared the screening rates according to HIV status and age group, before and after age 25 years. And before and after 2014, the year countries adopted the 2013 guidelines. We then used a data-driven optimal bandwidth selection procedure to estimate the guidelines’ average treatment effect (ATE), with a local polynomial regression discontinuity and robust bias-corrected confidence intervals. We validated the RDD methodology overall and for women with a significant ATE at the country-level analysis.

**Results:** We included 73179 women: 6680 (9.1%) living with HIV, 4328 (5.9%) with unknown HIV status, and 62171 (85.0) with a negative HIV status. 5726 (7.9%) reported having ever been screened; 4022 (6.5%) with unknown HIV status, 525 (12.1%) with a negative HIV status and 1179 (17.7%) living with HIV. Adolescent girls and young women living with HIV (AGYWLHIV) aged 15–24 reported screening less often (917 (13.7%)) than their peers with unknown (1677 (38.8%)) or positive HIV status (27278 (43.9%)) (P<0.001), or older women. The ATE of screening was 0 for women with unknown or positive HIV status, for whom the RDD was valid.

**Conclusion:** We found no evidence the 2013 WHO guidelines increased or reduced women’s cervical pre-cancer screening. However, AGYWLHIV reported screening less often. Policy makers should lower the age to start screening WLHIV from 25 to 15 to screen more AGYWLHIV. Studies are required to examine the impact of the guidelines on cervical pre-cancer screening in more countries.

## Introduction

Cervical cancer is the leading cause of death among women (1,2). According to recent global estimates, East and Southern African regions had the highest incidence of new cervical cancer cases across all regions (40 and 36 cases per 100,000 women years, respectively), and Africa had the highest mortality rates globally in 2020 (2). In 2013, the World Health Organization (WHO) issued comprehensive guidelines since 2010. It recommended cervical pre-cancer screening among women aged 30__49, and sexually active girls and women as soon as they test HIV-positive to prevent invasive cancer among women living with HIV (WLHIV). WLHIV are at higher risk of developing cervical cancer than women with an HIV-negative status (3). The guidelines indicated that the evidence for screening women with an HIV positive status to prevent cervical cancer was of lower quality than that for women with a negative or unknown HIV status (3). Many African countries (38 of 54) formulated cervical cancer prevention and treatment strategies based on the WHO guidelines (4). The WHO developed the guidelines from randomised controlled trials, systematic reviews, and expert guidance, among others (1,5). However, the impact of the 2013 WHO cervical pre-cancer screening and treatment guidelines and the age to start screening at the population level has not been studied.

Studies have examined the policy-level impacts of the WHO guidelines to expand the HIV treatment eligibility criteria and the “Treat all” guidelines to provide immediate anti-retroviral therapy (ART) to people living with HIV (PLHIV) regardless of their CD4 count (6,7,8). These studies found that the guidelines expanded the enrolment and retention of PLHIV in care (6,7,8). Research is required to identify whether the 2013 WHO cervical pre-cancer guidelines changed dynamics around screening for vulnerable women in east and southern Africa. The guidelines may have helped decision makers choose and implement the optimal screening and treatment strategy applicable in their context. Conversely, the new guidelines might have led decision makers to defer screening, for adolescent girls and young women living with HIV (AGYWLHIV) to age 30 and over. Examining whether trends in screening changed after the 2013 WHO guidelines may inform screening efforts for 2020–2030.

The WHO developed the 2013 guidelines to guide decision makers about which screening and treatment strategy to use in their settings. It recommended an overall screen-and-treat strategy with the treatment for precancerous lesions provided soon after a positive screening test result instead of after a sequence of tests and examinations. Besides nine recommendations, the guidelines contain a decision-making flow chart for choosing the best screen-and-treat strategy for a particular setting. It includes flow charts for women with a negative, unknown, or positive HIV status. It did not specify the age to start screening women with an HIV-positive status (3). The WHO later recommended starting screening WLHIV starting at age 25 (5). Stratifying analyses by HIV status and age groups, we examined the impact of the age, 25, to start screening on screening rates in Ethiopia, Malawi, Rwanda, Tanzania, Zambia, and Zimbabwe. We then used a regression discontinuity design (RDD) analysis to examine the impact of the WHO guidelines on women’s screening rates. The RDD methodology allows us to estimate whether the guidelines impacted screening (10).

## Methods

### Data sources

The Population-based HIV Impact Assessment (PHIA) survey data for countries in East and Southern Africa conducted between 2015 and 2018 which included questions on cervical cancer were used for this analysis. The countries and their respective survey periods are Ethiopia (2017__2018), Malawi (2015__2016), Rwanda (2018), Tanzania (2016__2017), Zambia (2016) and Zimbabwe (2015__2016). In these countries the women were asked if they were screened for cervical pre-cancer and if so, the last year screened, ranging from 2008 to 2018. The PHIA surveys aimed at evaluating the impact of HIV programmes in countries supported by the United States President’s Emergency Plan for AIDS Relief. The surveys collected a range of HIV biomarkers, and socio-demographic data among the respondents. The HIV biomarker data were confirmed with laboratory tests following the testing algorithm of the respective country. In each country the conduct of the survey was led by the Ministry of Health or an equivalent body. The ethical review board of the respective country reviewed and approved the survey protocol, including the informed consent procedures. The data collection followed ethical standards for conducting research on human subjects. All respondents provided informed verbal consent to participate in the survey based on the approved consent and assent procedures. The Head of the Household provided consent for household members to participate in the survey interviews. Then, adult household members aged 18 years and older provided consent to participate in the surveys, including for the HIV biomarker component. Children aged 10__17 were asked to participate in the survey only after their parents or guardians permitted their participation. The trained survey interviewers recorded the consent on the electronic tablets.

We combined each country’s household, adult individual and HIV biomarker data sets and pooled the data for the six countries. We restricted analyses to women aged 15__49 who had a negative, unknown, or positive HIV status and responded to the cervical pre-cancer screening questions. This group includes women aged 30__49 years, and WLHIV aged 25__49 for whom the WHO recommended screening for cervical pre-cancer (5). We identified, in the literature, that the countries’ cervical pre-cancer screening programmes were rule-based; they followed the WHO cervical pre-cancer screening recommendations before and after the 2013 WHO guidelines were issued, including the age to start screening women (4,11,12,13,14).

### Analysis

The primary outcome was self-reported ever being screened for cervical pre-cancer among all women, women with a negative, unknown, or HIV-positive status, between 2008 and 2018. We analysed the effect of the 2013 guidelines on screening rates. We considered women who reported being screened before 2014, as not having been exposed to the new guidelines, i.e. not screened based on the 2013 guidelines. Women who reported being screened in 2014, and after, were considered as having been exposed to the guidelines, presenting a sharp RDD analysis. In the sharp RDD analysis, at the cut-off, in this study the year 2014, the probability of exposure to the guidelines changes from 0 to 1, or from 1 to 0. Covariates included levels of education (no education, primary, secondary, and higher), household wealth status in wealth quintiles, rural-urban residence, ever being married (yes/no), age (15__24, 25__34, and 35__49), and country, for pooled analyses.

We coded ever being screened in binary form, indicating whether a woman had been screened, and the year of the last screening. We defined HIV status as negative, unknown if HIV test results were missing, or positive, from self-reported HIV status confirmed with HIV laboratory tests. The wealth quintiles assessed household wealth, ranking households based on household characteristics and asset ownership, from wealth quintile 1 (Q1) representing the poorest households to Q5 the wealthiest. The wealth quintile variable was provided in the dataset.

We described the sample with descriptive statistics and assessed differences in rates of screening for women aged 15__24, 25__34 and 35__49 years by HIV status with chi squared tests. We plotted the number of women screened by year last screened with a histogram for all women, and for women with a negative, unknown, or HIV-positive status to explore continuity in screening before and after 2014. We used the rdrobust software package for data-driven bandwidth selection (15). We selected the optimal bandwidths that minimized the mean squared errors of the regression discontinuity (MSERD) for both the pooled and the country level analyses, using a data-driven bandwidth selection procedure with the rdbwselect function. We fitted the selected bandwidth for each group of women (10). Using the rdrobust function, we estimated the magnitude of the discontinuity in screening in 2014 with a local polynomial regression discontinuity with robust bias-corrected 95% confidence intervals. In the optimal bandwidth area, linearity of the data is assumed, and estimated the average treatment effect (ATE) of the intervention (i.e., the 2013 WHO guidelines) with a local linear polynomial regression. In the estimation, we adjusted for covariates. Standard errors were clustered at the country__level for the pooled analysis, and the third nearest neighbour (nn 3) for country__level analyses. The third nearest neighbour cluster was an option for the country-level analysis.

We calculated the local average treatment effect (LATE) and the treatment effective derivative (TED) for sensitivity analyses. The LATE presents the treatment effect in the local area of the linear estimation. Its magnitude should be close to the ATE. The TED estimates the stability of the effect in the local area of the linear estimation. If its p-value is greater than 0.05, the effect is considered stable in the local area of the linear estimation and the RDD is validated.

We conducted several tests to validate the RDD methodology for all groups of women in the pooled analyses, and only for the countries with groups of women with a significant ATE and relative change in ATE. The tests included the alternative discontinuity thresholds, checking the balance in pre-determined covariates at the threshold and no sorting at the cut-off (Supplementary Table 2). Failing to reject the null hypothesis of the RDD tests at p = 0.05 i.e., the p-values of the RDD test were greater than 0.05, validated the assumption of the RDD tested (16,17). The validity of the RDD assumptions attributed the changes in screening rates observed in the analyses to the guidelines. We used Stata version 14.0 for the analyses. This study did not require ethical clearance because the data is de-identified and publicly available at https://phia-data.icap.columbia.edu.

## Results

Table 1 shows the sample, comprising 73179 women, 6680 (9.1%) with a positive, 4328 (5.9%) with an unknown and 62171 (85.0%) with a negative HIV status. The samples of women per country ranged from 10157 in Malawi to 15455 in Tanzania. WLHIV were generally older than women with an unknown or a negative HIV status. Self-reported screening rates were higher in all surveyed countries among WHLHIV (17.7%, n = 1179) than among women with an unknown (12.1%, n = 525) or negative HIV status (6.5%, n = 4022) (p<0.001). The screening rates ranged from 10.2% (n = 50) and 6.6% (n = 930) among WLHIV or women with a negative HIV status in Rwanda to 22.5% (n = 342), 14.9% (n = 155) and 9.6% (n = 816) among WLHIV, women with unknown, or negative HIV status in Zambia.

**Table 1:**
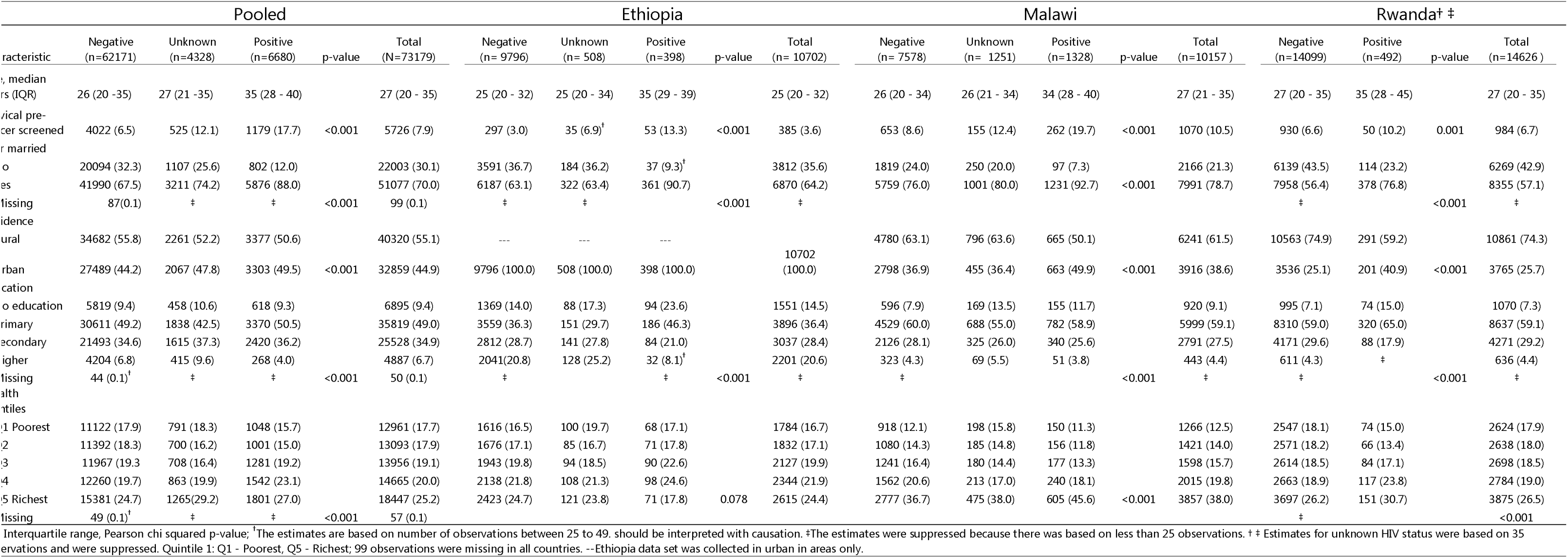
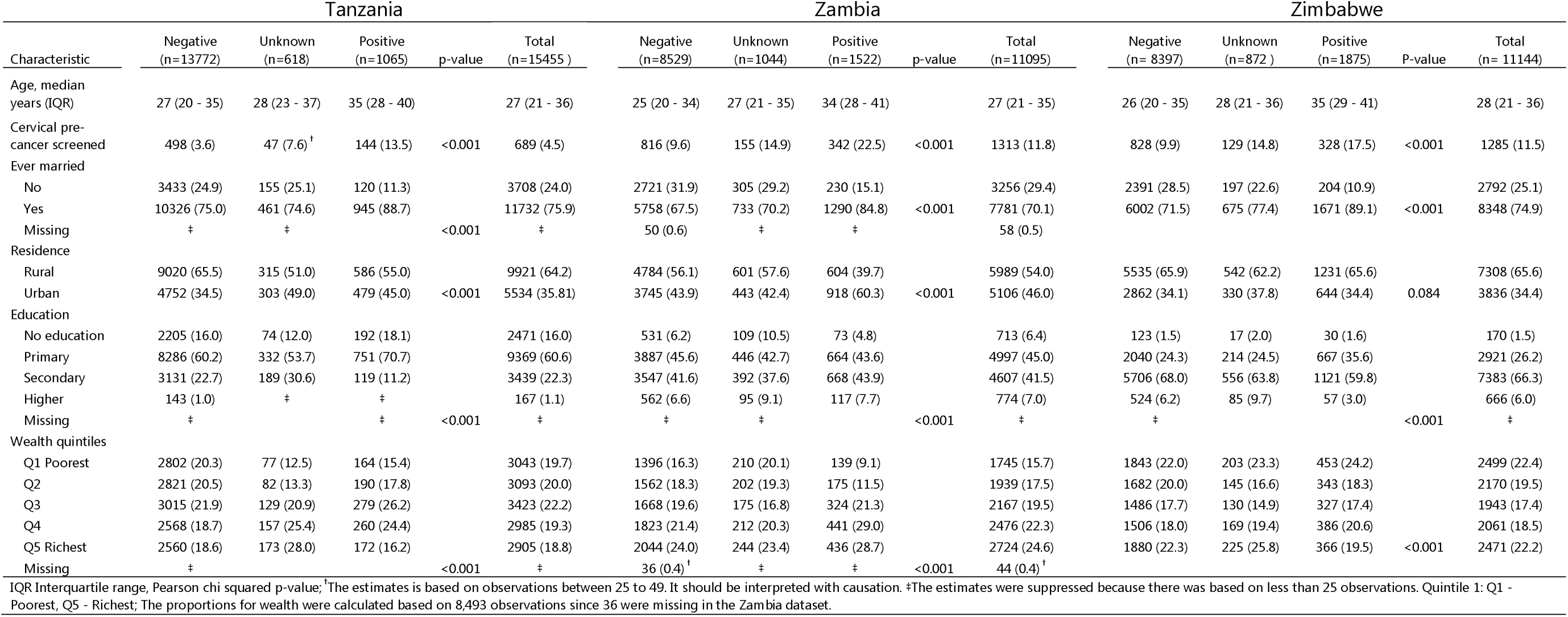
Baseline characteristics and self-reported cervical pre-cancer screening rates among women by country and HIV status, presented as medians with IQR, and frequencies with rcentages (PHIA 2015L18).

Over two-thirds of women were ever married in all surveyed countries. More than half resided in rural areas except in Ethiopia, where the survey was conducted in urban areas. Less than half of women had primary level education in Ethiopia, Zambia, and Zimbabwe, whereas more than half had primary level of education in Malawi, Rwanda, and Tanzania. More women were from households in the wealth quintile 5, except for Tanzania (Table 1).

Adolescent girls and young women living with HIV (AGYWLHIV) aged 15LJ24 reported screening rates that were at least two times lower than their peers with a negative or unknown HIV status in all countries. In contrast, adolescent girls and young women (AGYW) aged 15LJ24 with a negative or unknown HIV status reported higher screening rates than women aged 25LJ34 or 35LJ49 with a negative or unknown HIV status (Figure 1). The differences were significant (Supplementary Table 1).

**Figure 1:**
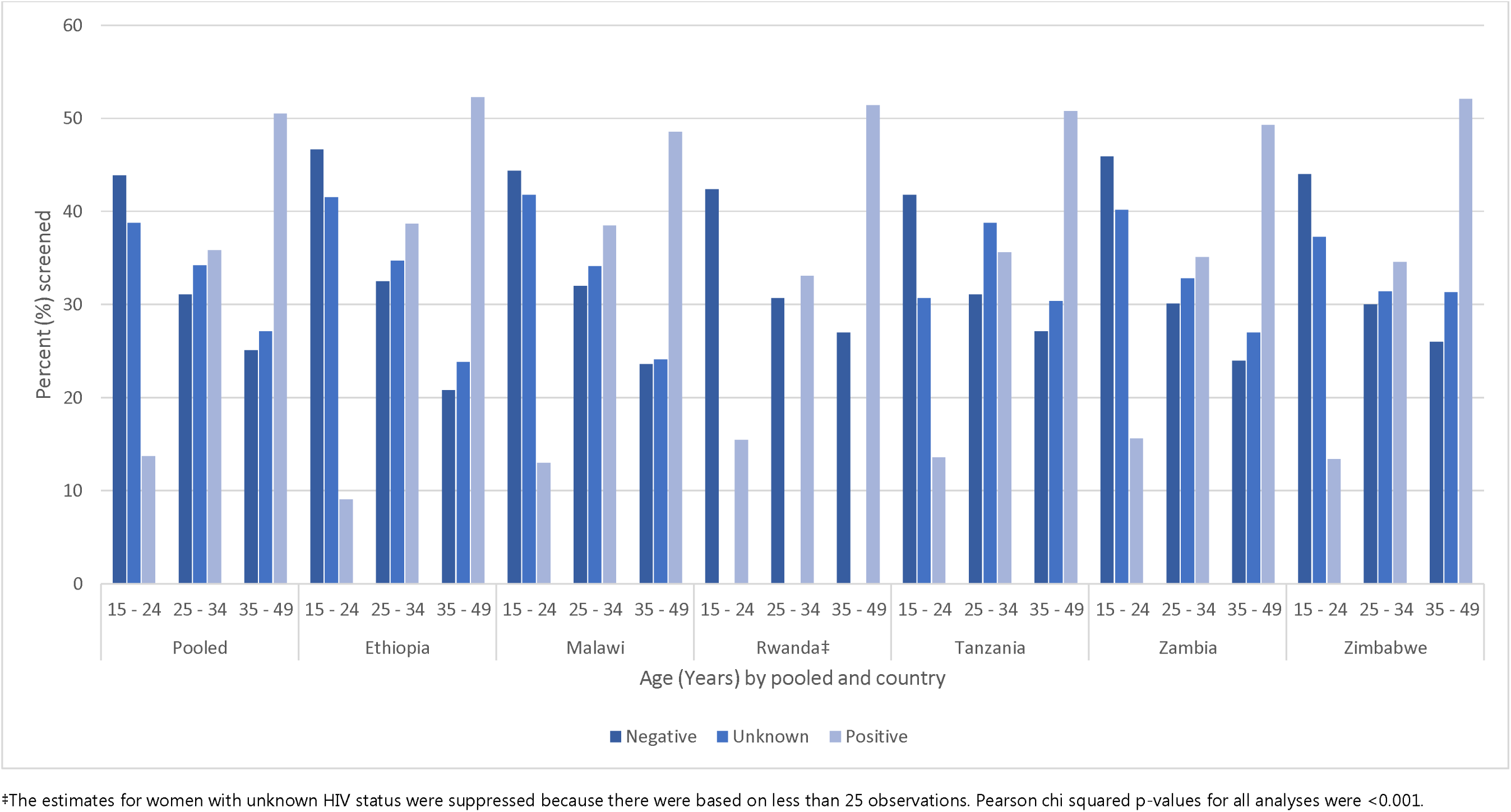
Self-reported cervical pre-cancer screening rates among women by country, HIV status, and age group, presented as percentages (PHIA 2015L18).

Figure 2 shows the number of women screened for cervical pre-cancer in all six countries by HIV status, and year last screened. The period of women’s screening ranged from 2008 to 2018. Beside Rwanda, all countries surveyed had data before and after 2014. Rwanda had data for women with unknown HIV status after 2014, which were excluded from the RDD estimation.

**Figure 2:**
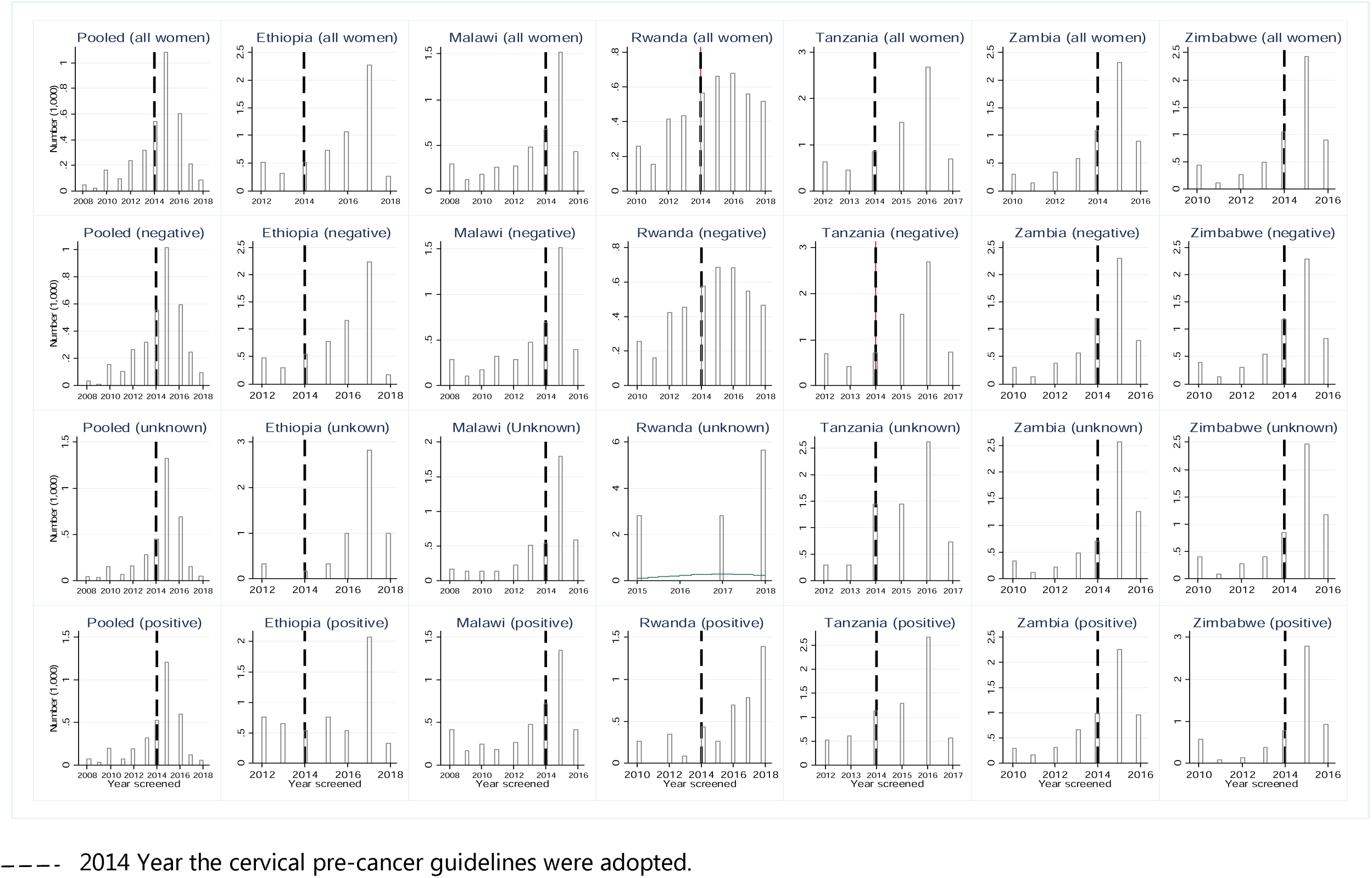
Number of self-reported screened for cervical pre-cancer among women stratified by country, HIV status and year screened (PHIA 2015L18).

Table 2 shows no evidence the guidelines increased or reduced the screening rates in the analyses overall stratified by the women’s HIV status, or the country-level (Table 2), or by age group (results not presented). The ATE was small and less than 0.001% in the pooled analysis. In the country-level analysis, the ATE was small (<0.001%) for all women, and for women with a negative or a positive HIV status in Malawi, Rwanda, and Zambia (Table 2). The RDD was valid. The assumptions held for implementing the RDD method, for women with unknown or HIV-positive status for pooled analysis; not for all women or women with an HIV-negative status. Although the related change in ATE for WLHIV in Ethiopia and Tanzania were significant, the RDD was not valid (Supplementary Table 2).

**Table 2:**
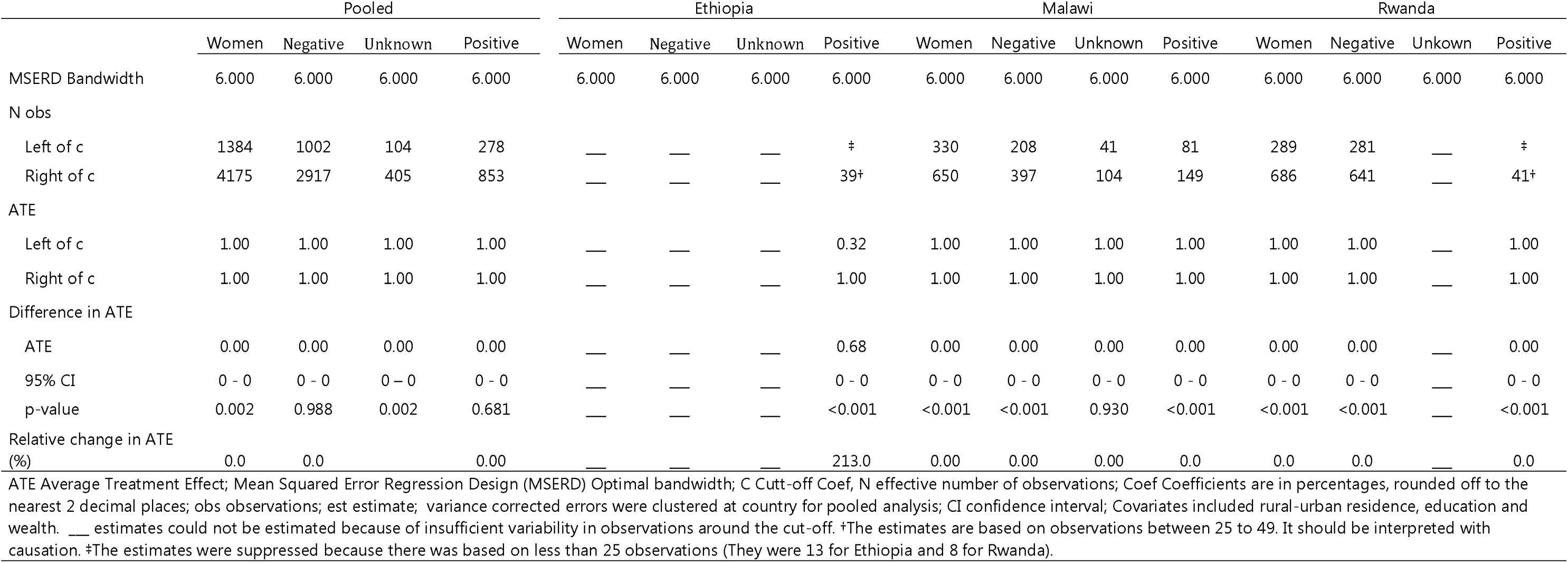
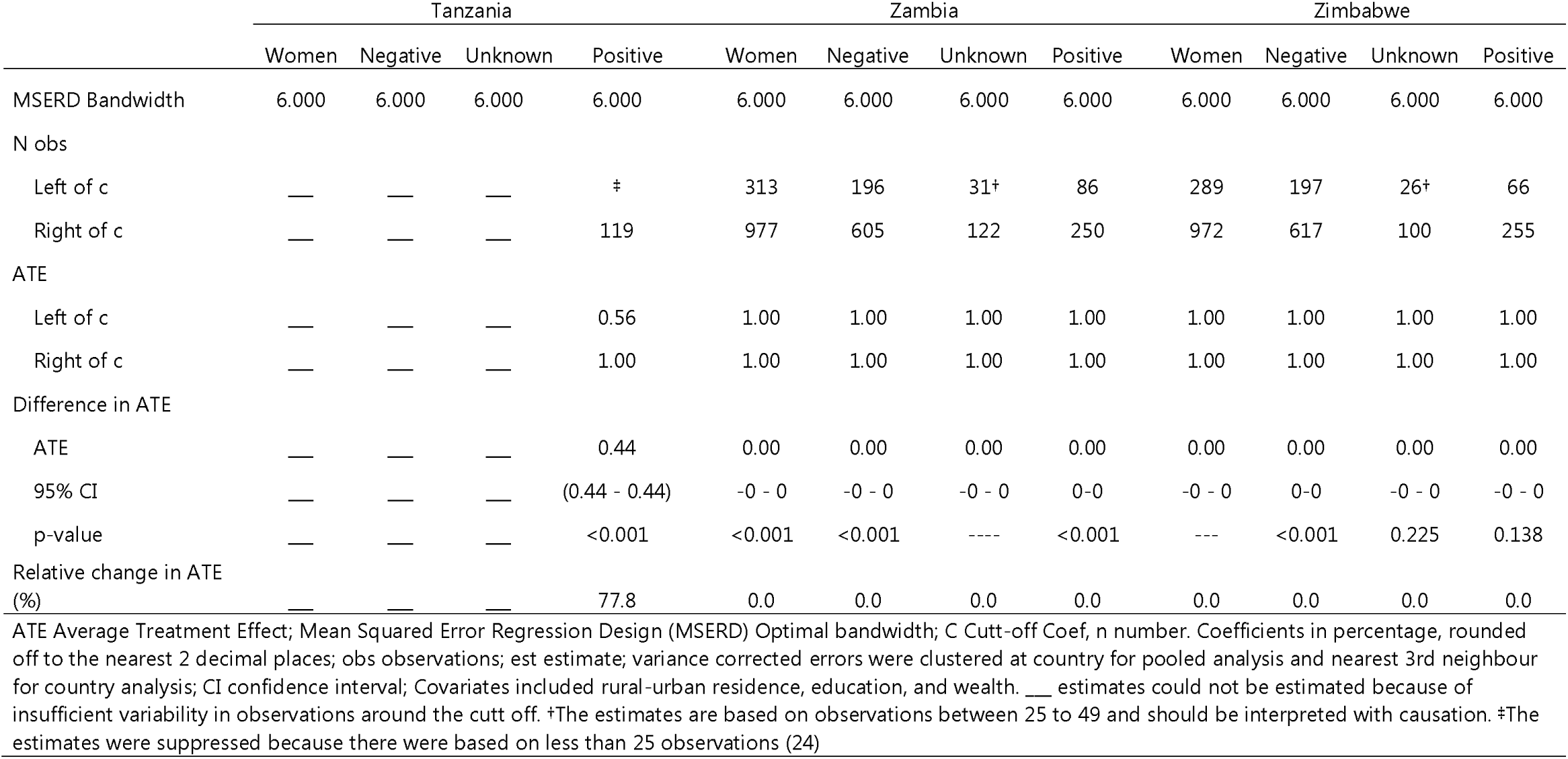
Effect of the 2013 WHO cervical precancer guidelines on women’s screening rates stratified by pooled and country analyses (PHIA 2015 18) (Local polynormal <u>point robust estimate).</u>

## Discussion

This study examined the impact of the 2013 WHO guidelines for screening and treating cervical precancerous lesions for cervical cancer prevention on women’s screening rates in six East and Southern African countries. We found no evidence that the guidelines negatively affected women’s screening rates. However, AGWLHIV reported screening less often than their peers with a negative, or unknown HIV status, or older women. The age to start screening WLHIV should be lowered from 25 to 15 to screen more AGYWLHIV aged 15_24. Countries should adopt, implement, and monitor the impact of the guidelines on screening.

Our main finding was that the 2013 WHO cervical pre-cancer guidelines did not reduce or increase women’s screening rates. This result applied to women with unknown or HIV-positive status in pooled analyses. Our result supports the evidence that shows that the recommendations often have heterogeneous impacts on objectives. The guidelines sometimes have unintended consequences (6,7,8,16,17,18). In the United States of America (USA), state insurance mandates to cover costs for Pap smear tests, increased Pap testing rates by 1.3 percentage points among women aged 19_64. The impacts were more pronounced in black and Latino women than in white women (18). In sub–Saharan Africa, the recommendations to expand ART eligibility increased ART initiation and retention in care with greater impacts on PLHIV who met or nearly met the eligibility criteria to access ART (6,7). The recommendation did not prevent people with advanced HIV disease from initiating ART (6,7). However, one study found that the “Treat all” guidelines may have reduced pre-ART CD4 monitoring, weakening advanced HIV disease management (8).

One reason for our findings is that several countries began their cervical cancer prevention programmes around 2008, with government support, followed by a rapid scale-up. Adopting and implementing the new guidelines might have not imposed significant resource reallocations in countries’ cervical pre-cancer screening programmes. The 2013 guidelines simplified the screening and treatment process, encouraging decision-makers to use the screening and treatment strategy suitable to their context. Further comprehensive cervical cancer control guidelines were launched in December 2014, providing additional support and guidance to decision-makers and stakeholders in accelerating cervical cancer screening and treatment of pre-cancerous lesions (19). Altogether, the WHO established the 2013 cervical pre-cancer screening guidelines accompanied by deliberate efforts, including training, and media outreach, to support decision-makers in countries in adopting and implementing the guidelines (3). This result suggests that the guidelines and the efforts surrounding their dissemination are crucial in ensuring decision-makers effectively manage screening programmes and prevent unintended consequences, such as, reducing screening for vulnerable women.

We found that AGYWLHIV reported a screening rate that was at least two times lower than that of their peers with a negative or unknown HIV test result, or older women. Their peers with a negative or unknown HIV status reported higher screening rates than older women with a negative or unknown HIV status. This result reveals a systematic exclusion of AGYWLHIV from screening, an intended result of implementing the 2013 WHO guidelines. Although the guidelines recommended screening sexually active girls and women immediately after they tested HIV-positive, decision-makers might not have prioritized screening AGYWLHIV. They might have only screened them if AGYWLHIV or their providers requested the screening (11,15). AGYWLHIV might have not been aware of their HIV-positive status and less likely to access ART, and associated services including cervical cancer screening than older WLHIV (20,21). Further, AGYWLHIV might have not accessed ART and screening because of stigma and discrimination associated with HIV. In this study, we found higher screening rates among older WLHIV than any group of women. Policy makers might have prioritised screening older WLHIV than AGYWLHIV or other sub-groups of women, showing that the countries followed the WHO recommendations.

However, the potential risk to cervical cancer for AGYWLHIV in East and Southern Africa is worth re-examining. AGYW aged 15_24 are vulnerable. They accounted for 81% of the 250,000 new HIV infections among young people in 2021 (22). AGWYLHIV may be at higher risk of human papillomavirus (HPV) infection, and cervical cancer than their peers without HIV (23). HPV takes longer to clear in AGYWLHIV than among their peers without HIV (24). More AGYWLHIV in East and Southern Africa could be screened by lowering the age to start screening WLHIV. In the USA, stake holders proposed lowering the age to start screening from 25 to 21 (25). One reason for this proposal is that cervical cancer is more prevalent among women aged 25_29 in the USA. Screening women younger than 25 would reduce the risk of cervical cancer in women aged 25_29 (25). A further reason is the lack of national cancer screening programmes to screen and detect cervical pre-cancer lesions among young women (25). The countries in our study do not have large scale comprehensive national screening programmes that reach a diverse group of women at risk of cervical cancer (4,11). The WHO included guidance for screening WLHIV aged 25_49 years in the 2021 guidelines (1,5). The 2021 guidelines excluded AGYWLHIV from screening, including those born with HIV, who may have an elevated risk of cervical cancer. It argued that the evidence on the utility of screening AGYWLHIV was limited and that WLHIV face more screening over their life with increased potential harm from multiple screenings (23). Lowering the age to start screening WLHIV to include 15_24 years-olds, and prioritizing AGWYLHIV in screening, may lead to screening younger women who do not need screening (25). The potential for over-screening and its associated risks may be justified in East and Southern Africa where screening rates are low and the risk to cervical cancer among AGWLHIV is high.

Our study has several limitations and strengths. Women may have recalled the responses inaccurately because they are self-reported. Countries are flexible in adapting the guidelines to their context (19). However, they may align their screening programmes to the WHO screening guidance. The results are valid for the respondents in the local areas of the regression, typical of RDD analyses. Although the RDD assumptions for this study were valid for women with unknown or positive HIV status in pooled stratified analyses, the sample size was small for several variables reducing the variability of the observations for the estimations. A strength of this study is that we chose the optimal bandwidths apriori using data-driven bandwidth selection procedure, preventing us from selecting biased bandwidths. Another strength is that we used quality population-based data with high response rates. The assumptions of the RDD were valid for women with unknown or HIV positive status. Therefore, we could make causal inference of changes in women’s screening rates based on the 2013 WHO guidelines for these group of women.

## Conclusion

We found no evidence that the 2013 WHO guidelines for screening and treating pre- cancerous lesions for the prevention and treatment of cervical cancer reduced or increased women’s screening rates. However, AGYWLHIV reported screening less often than their peers or older women. Policy makers may need to lower the age to start screening women living with HIV from 25 to 15 years to screen more AGWYLHIV. Studies are required to examine the impact of the guidelines including the age to start screening women living with HIV on cervical cancer screening to screen more vulnerable AGYWLHIV.

## Data Availability

The data is publicly available. It can be accessed with a request at https://phia-data.icap.columbia.edu

## Acknowledgements

We are grateful for the helpful and comprehensive comments from Amber Peterman and the Joint United Nations Programme on HIV/AIDS (UNAIDS). Olivia Keiser was supported by the Swiss National Science Foundation (grant no 202660). The funders or UNAIDS had no role in study design, data collection and analysis, publication decision or manuscript preparation. The views in this article are solely those of the authors and not UNAIDS’.

## Authors’ contributions

Conceptualization, DC. Methodology, DC. Validation, OK, JE, SA, AN, CO, JB, AS and OK. Formal analysis, DC, JE, SA, CO, JB, AN, AS and OK; data curation, DC writing—original draft preparation, DC.; writing—review and editing, DC, OK, JE, SA, AN, CO, JB, AS and OK; visualization, DC. All authors have read and agreed to the published version of the manuscript.

## Ethics declaration

This study did not require ethical review because the personal details in the data are de-identified, and the data is publicly available. It can be accessed with a request at https://phia-data.icap.columbia.edu

## Bibliography

1. World Health Organization. WHO guideline for screening and treatment of cervical pre-cancer lesions for cervical cancer prevention, second edition. Geneva: : World Health Organization; 2021. Report No.: 978 92 4 0040434.

2. Singh D, Vignat J, Lorenzoni V, Eslahi M, Ginsburg O, Lauby-Se B. Initiative, Global estimates of incidence and mortality of cervical cancer in 2020: a baseline analysis of the WHO Global Cervical Cancer Elimination. Lancet Glob Health. 2023;(10.1016/S2214-109X(22)00501-0): 11: e197–206.

3. World Health Organization. WHO guidelines for screening and treatment of precancerous lesions for cervical cancer prevention. Geneva: World Health Organization; 2013. Report No.: ISBN 978 92 4 154869 4.

4. Akanda R, Kawale P, Moucheraud C. Cervical cancer prevention in Africa: A policy analysis. Journal of cancer policy. 2022 Jan; 32(Doi.org/10.1016/j.jcpo.2021.100321).

5. World Health Organization. Global strategy to accelerate the elimination of cervical cancer as a public health problem. Geneva: World Health Organization; 2020. Report No.: ISBN 978-92-4- 001410-7.

6. Bor J, Fox MP, Rosen S, Venkataramani A, Tanser F, Pillay D, et al. Treatment eligibility and retention in clinical HIV care: A regression discontinuity study in South Africa. Plos Medicine. 2017 Nov;(Doi.org/10.1371/journal.pmed.1002463).

7. Mody A, Sikazwe I, Czaicki NL, Mwanza MW, Savory T, Sikombe K, et al. Estimating the real-world effects of expanding antiretroviral treatment eligibility: Evidence from a regression discontinuity analysis in Zambia. Plos medicine. 2018 June;(Doi.org/10.1371/journal.pmed.1002574).

8. Zaniewski E, Brazier E, Ostinelli CHD, Wood R, Osler M, Technau KG, et al. Regression discontinuity analysis demonstrated varied effect of Treat-All on CD4 testing among Southern African countries. Journal of Clinical Epidemiology. 2021; 140(Doi.org/10.1016/j.jclinepi.2021.09.001.): 101–110.

9. Stelzle D, Tanaka LF, Lee KK, Khalil AI, Baussano I, Shah ASV, et al. Estimates of the global burden of cervical cancer associated with HIV. Lancet Glob Health. 2020 Nove;(10.1016/S2214-109X(20)30459-9).

10. Lee DS, Lemieux T. Journal of Economic Literature. 2010 June; 48(http://www.aeaweb.org/articles.php?doi=10.1257/jel.48.2.281): 281–355.

11. Asangbeh-Kerman SL, Davidović M, Taghavi K, Kachingwe J, Rammipi KM, Muzingwani L, et al. Cervical cancer prevention in countries with the highest HIV prevalence: a review of policies. BMC Public Health. 2022 Aug; 22(1) Doi: 10.1186/s12889-022-13827-0.): 1530.

12. Ethiopia FDRo. Guideline for cervical cancer prevention and control in Ethiopia. Addis Ababa: Federal Democratic Republic of Ethiopia; 2015.

13. The Government of Malawi Ministry of Health UNPF (. National cervical cancer control strategy 2016 - 2020. The Government of Malawi, Ministry of Health; 2017.

14. Binagwaho A, Ngabo F, Wagner CM, Gatera M, Nutt CT, Nsanzimana S. Integration of comprehensive women’s health programmes into health. Bull World Health Organ. 2013 May; 91(Doi.org/10.2471/BLT.12.116087): 697–703.

15. Pry JM, Manasyan A, Kapambwe S, Taghavi K, Duran-Frigola M, Mwanahamuntu M, et al. Cervical cancer screening outcomes in Zambia, 2010-19: a cohort study. Lancet Glob Health. 2021 Jun ; 9(6) doi: 10.1016/S2214-109X(21)00062-0.): e832–e840.

16. Carpenter C, Dobkin C. The Effect of Alcohol Consumption on Mortality: Regression Discontinuity Evidence from the Minimum Drinking Age. American Economic Journal: Applied Economics. 2009 Jan; 1(1): 164–8.

17. Kadiyala S, Strump E. How Effective is Population-Based Cancer Screening? Regression Discontinuity Estimates from the US Guideline Screening Initiation Ages. Forum for Health Economics and Policy. 2016; 19(1) Doi.org/10.1515/fhep-2014-0014): 87–139..

18. Bittler MP, Carpenter CS. Effects of state cervical cancer insurance mandes on pap test rates. Health services research. 2017; 52 (1)(Doi:10.1111/1475-6773.12477).

19. World Health Organization. Comprehensive cervical cancer control A guide to essential practice - second edition. Geneva: World Health Organization; 2014. Report No.: ISBN 978 92 4 154895 3.

20. Haas AD, Radin E, Hakim AJ, Jahn A, Philip NM, Jonnalagadda S, et al. Prevalence of nonsuppressed viral load and associated factors among HIV-positive adults receiving antiretroviral therapy in Eswatini, Lesotho, Malawi, Zambia and Zimbabwe (2015 to 2017): results from population-based nationally representative surveys. J Int AIDS Soc. 2020 Nov; 23(11)(Doi: 10.1002/jia2.25631): e25631.

21. Chipanta D, Amo-Agyei S, Giovenco D, Estill J, Keiser O. Socioeconomic inequalities in the 90–90– 90 target, among people living with HIV in 12 sub-Saharan African countries — Implications for achieving the 95–95–95 target — Analysis of population-based surveys. eClinicalMedicine. 2022 September;(Doi.org/10.1016/j.eclinm.2022.101652).

22. Joint United Nations Programme on HIV/AIDS S (UNAIDS). Dangerous inequalities: World AIDS Day report. Geneva: Joint United Nations Programme on HIV/AIDS S (UNAIDS); 2022. Report No.: UNAIDS/JC3068E.

23. Dhokotera T, Bohlius J, Egger M, Spoerri A, Ncayiyana JR, Naidu G, et al. Cancer in HIV-positive and HIV-negative adolescents and young adults in South Africa: a cross-sectional study. BMJ Open. 2021; 11(10): e043941. 2021 Oct; 11 (10)(Doi: 10.1136/bmjopen-2020-043941): e043941.

24. Moscicki AB, Ellenberg JH, Farhat S, Xu J. Persistence of Human Papillomavirus Infection in HIV-Infected and -Uninfected Adolescent Girls: Risk Factors and Differences, by Phylogenetic Type. The Journal of Infectious Diseases. 2004 July; 190(1): 37–45.

25. Moscicki AB, Perkins RB, Saville M, Brotherton JM. Should cervical cancer screening be performed before the age of 25 years? J Low Genit Tract Dis. 2018 Oct;(Doi:10.1097/LGT.0000000000000434): 348–351.

